# Quantitative bias analysis for mismeasured variables in health research: a review of software tools

**DOI:** 10.1101/2024.12.12.24318922

**Authors:** Codie J.C. Wood, Kate Tilling, Jonathan W. Bartlett, Rachael A. Hughes

## Abstract

**Background:** Measurement error and misclassification can cause bias or loss of power in epidemiological studies. Software performing quantitative bias analysis (QBA) to assess the sensitivity of results to mismeasurement are available. However QBA is still not commonly used in practice, partly due to a lack of knowledge about these software implementations. The features and particular use cases of these tools have not been systematically evaluated.

**Methods:** We reviewed and summarised the latest available software tools for QBA in relation to mismeasured variables in health research. We searched an electronic database, Web of Science, for studies published between **1**^st^ January 2014 and **1**^st^ May 2024 (inclusive). We included epidemiological studies that described the use of software tools for QBA in relation to mismeasurement. We also searched for tools catalogued on the CRAN archive, Stata manuals, and Stata’s *net* command available from within Stata or from the IDEAS/RePEc database. Tools were included if they were purpose-built, had documentation and were applicable to epidemiological research. Data on the tools’ features and use cases was then extracted from the full article texts and software documentation.

**Results:** 17 publicly available software tools for QBA were identified, accessible via R, Stata, and online web tools. The tools cover various types of analysis, including regression, contingency tables, mediation analysis, longitudinal analysis, survival analysis and instrumental variable analysis. However, there is a lack of software tools performing QBA for misclassification of categorical variables and measurement error outside of the classical model. Additionally, the existing tools often require specialist knowledge.

**Conclusions:** Despite the availability of several software tools, there are still gaps in the existing collection of tools that need to be addressed to enable wider usage of QBA in epidemiological studies. Efforts should be made to create new tools to assess multiple mismeasurement scenarios simultaneously, and also to increase the clarity of documentation for existing tools, and provide tutorials and examples for their usage. By doing so, the uptake of QBA techniques in epidemiology can be improved, leading to more accurate and reliable research findings.

## 1 Introduction

In epidemiological and population health studies, we often aim to estimate the causal effect of an exposure or treatment on an outcome (referred to as the exposure effect) while adjusting for confounders or other variables [1]. Most methods of estimating an exposure effect rely on the assumption that sufficient confounders are known and have been included in the model, and that included variables have been measured without error. When data are obtained for epidemiological studies, there is potential for some of the variables to be measured with error and so this assumption may not be plausible, though this is often unacknowledged [2]. Where we have categorical or binary variables measured with error (as opposed to continuous variables), we refer to measurement error as misclassification. Throughout, the umbrella term “mismeasurement” is used to capture both scenarios.

It is a common misconception that mismeasured variables will always bias the effect estimate towards the null [3]. In fact, the impact of mismeasurement on an effect estimate depends on a number of factors, including the role of the variable(s) in which the mismeasurement occurs (i.e. if it is the outcome, exposure or other covariate), the type of the variable (i.e. if it is binary, continuous, or categorical)[4, 5], if errors in multiple variables are dependent on each other [6], the type of analysis being conducted, and whether the mismeasurement is differential (i.e. some aspect of the error distribution depends on another variable) [7].

Failing to account for mismeasurement can result in problems such as decreased statistical power, biased effect estimates (either towards or away from the null), and inaccurate representations of estimate uncertainty [8]. Any of these issues could result in the reporting of erroneous study conclusions. As such, it is important to account for and quantify the potential effects of mismeasurement within a study, as study results can inform government policies and development of large-scale health interventions. Despite this, most medical publications make no adjustment for and have little discussion of how mismeasurement potentially impacts results [9].

There exist many methods to adjust for mismeasurement, which have been described extensively in the literature [7, 8, 10]. These methods typically require some form of ancillary data, such as validation data (either internal or external), or replication data [8]. However, ancillary data is often not readily available. In these cases, sensitivity analyses such as a quantitative bias analysis (QBA) can be used to evaluate the potential impact of mismeasurement on a study’s conclusions.

QBA consists of a group of statistical methods for assessing uncertainty arising due to biases in a study [11]. It can be applied to various biases, including unmeasured confounding [12] and selection bias [13]. Here, we focus on QBA for mis-measurement, which is used to quantify the potential impact of mismeasurement or to assess how severe it would need to be to change a study’s conclusions. This allows analysts to assess the robustness of the study conclusions to the assumption of no mismeasurement. See Section 1.1 for further information on QBA.

Currently, QBA methods are not employed as a standard practice. A recent review found that QBA usage in epidemiology increased between 2006 and 2019 [13], but it was still relatively rare. This is partly attributed to the lack of available software for implementing these analyses, as well as limited awareness about such tools and lack of adequate teaching to epidemiological researchers about QBA [1, 14].

There have been several reviews of implementations of QBA methods for unmeasured confounding, misclassification and selection bias in epidemiology and health-related fields [13, 15–17]; however, these papers did not review software implementations of the methods they discussed. Software implementations of QBA methods were reviewed in [12]; however, this review was restricted to bias caused by unmeasured confounding.

In this scoping review we aim to identify the latest available software tools that implement a QBA for mismeasurement within epidemiological studies quantifying an exposure effect estimate, and provide details on their features and use cases. This will increase awareness of the software and, alongside developments in guidance for researchers on appropriate QBA implementations [15, 16, 18], promote its usage as standard practice for health research. We also aim to highlight potential future areas for software development.

### 1.1 Background on quantitative bias analysis

A QBA for mismeasurement quantifies the likely magnitude and direction of the bias under different plausible assumptions about the mismeasurement process (assuming no other sources of bias). Generally, a QBA requires a model (known as a bias model) for the observed data and the mismeasurement errors [11]. The bias model includes one or more parameters (known as bias or sensitivity parameters), which cannot be estimated from the observed data. These bias parameters encode the user’s assumptions about the mismeasurement process, determining the magnitude and direction of the bias-adjustment. For example, in the case of misclassification, the bias parameters may include some combination of the sensitivity, specificity, positive predictive value and negative predictive values. For a continuous variable measured with error, bias parameters may include reliability values or error magnitudes. Since the observed data do not provide enough information to estimate the bias parameters, the user must pre-specify information about the values of the bias parameters to enable estimation of the remaining parameters of the bias model and obtain an estimate of the parameter of interest (e.g., exposure effect) adjusted for bias due to mismeasurement. This information is usually obtained from external sources such as validation studies, published literature, data constraints or expert opinion [18].

QBA methods can broadly be classified into two categories: deterministic and probabilistic [11]. A deterministic QBA specifies one or more values for each bias parameter. A “simple bias analysis” fixes each bias parameter to a single value (i.e., treating the bias parameter values as known) and outputs a single bias-adjusted estimate of the exposure effect [11]. Typically, the bias parameters are unknown and so the analyst will need to perform a “multidimensional bias analysis” where multiple values are specified for each bias parameter, and the bias model is repeatedly fitted for each combination of bias parameter values. For example, in the case of multidimensional bias analysis for misclassification, we could consider different pairs of sensitivity and specificity values. A multidimensional bias analysis then outputs multiple bias-adjusted estimates. When there is very limited information about plausible values for the bias parameters, a tipping point analysis can be conducted to explore which combinations of values of the bias parameters would overturn study conclusions.

In a probabilistic QBA, the analyst specifies a prior probability distribution for the bias parameters. Using this prior distribution, the analyst can specify information about the range of plausible values of the bias parameters, the value combinations that are most likely to occur, and the analyst’s uncertainty about this information. A probabilistic QBA generates an empirical distribution of bias-adjusted estimates, which can then be summarised to give a point estimate (i.e., the most likely bias-adjusted estimate under the QBA’s assumptions) and an interval estimate (i.e., defined to contain the true exposure effect with a specific probability).

Two main approaches to a probabilistic QBA are Bayesian bias analysis (where the prior distribution of the bias parameters is combined with the likelihood function for the data) and Monte Carlo bias analysis (where values of the bias parameters are directly sampled from their prior distribution and then used to fix the bias parameters to enable estimation of the bias-adjusted exposure effect) [19].

A multiple QBA (also known as a multiple-bias analysis) assesses the sensitivity of study results to multiple sources of bias such as mismeasurement, unmeasured confounding, and selection bias. A sequential multiple QBA adjusts for one bias at a time, where the order of adjustment should be based on the reverse order in which the biases occurred during the data generation process [11]. A downside of the sequential approach is that the final bias-adjusted estimate may differ depending on the order of adjustment [20]. A simultaneous multiple QBA avoids this issue because it adjusts for the multiple sources of bias simultaneously [20].

## 2 Methods

We searched for QBA software described in health research articles, as well as those available in software databases, published between 1^st^ January 2014 and 1^st^ May 2024 (inclusive). We define a QBA as a method that adjusts for mismeasurement using a model that includes one or more bias parameters. Also, “software” is defined as a web tool, package, or code that is publicly available to use, is not specific to a particular data example, and is accompanied by documentation. To be classified as having “documentation”, a tool must provide enough information for users to understand its function and implementation without reliance on an external publication. This includes a user guide or in-code comments that explain the syntax, input requirements, and expected outputs. Examples of tools not meeting our software definition would be tools that were not publicly available, code files for specific examples that the user had to manually edit to apply to their study, and software that lacked documentation describing how to use the code, such as raw code files without explanatory comments. We searched published literature from health research and software databases, as this is how most health researchers would identify methods and tools they can use.

We did not look for methods within textbooks as these are not publicly available and often cannot be easily searched by applied researchers.

This review was written following the Preferred Reporting Items for Systematic reviews and Meta-Analyses extension for Scoping Reviews (PRISMA-ScR) guidelines [21], and the PRISMA-ScR checklist can be found in Additional File 1. Our search was conducted in three steps: search implementation, eligibility screening and data extraction.

### 2.1 Publication search

In our first step, we used Web of Science to identify papers that mentioned all of the terms “measurement error”, “bias analysis” and “software” (or some other synonym of these terms) in either the title, abstract or as keywords. The specific search terms used can be seen in Figure 1 [22]. The search was applied to databases “Web of Science Core Collection”, “BIOSIS Citation Index”, “KCI-Korean Journal Database”, “MEDLINE” and “SciELO Citation Index”. We excluded from our search any meeting abstracts, clinical trials or patents. In addition, we excluded any articles published in journals outside of the fields of statistics, medicine, population health, and epidemiology and so deemed out of the scope of health research. A list of the excluded journals is given in Figure 2.

**Fig. 1.**
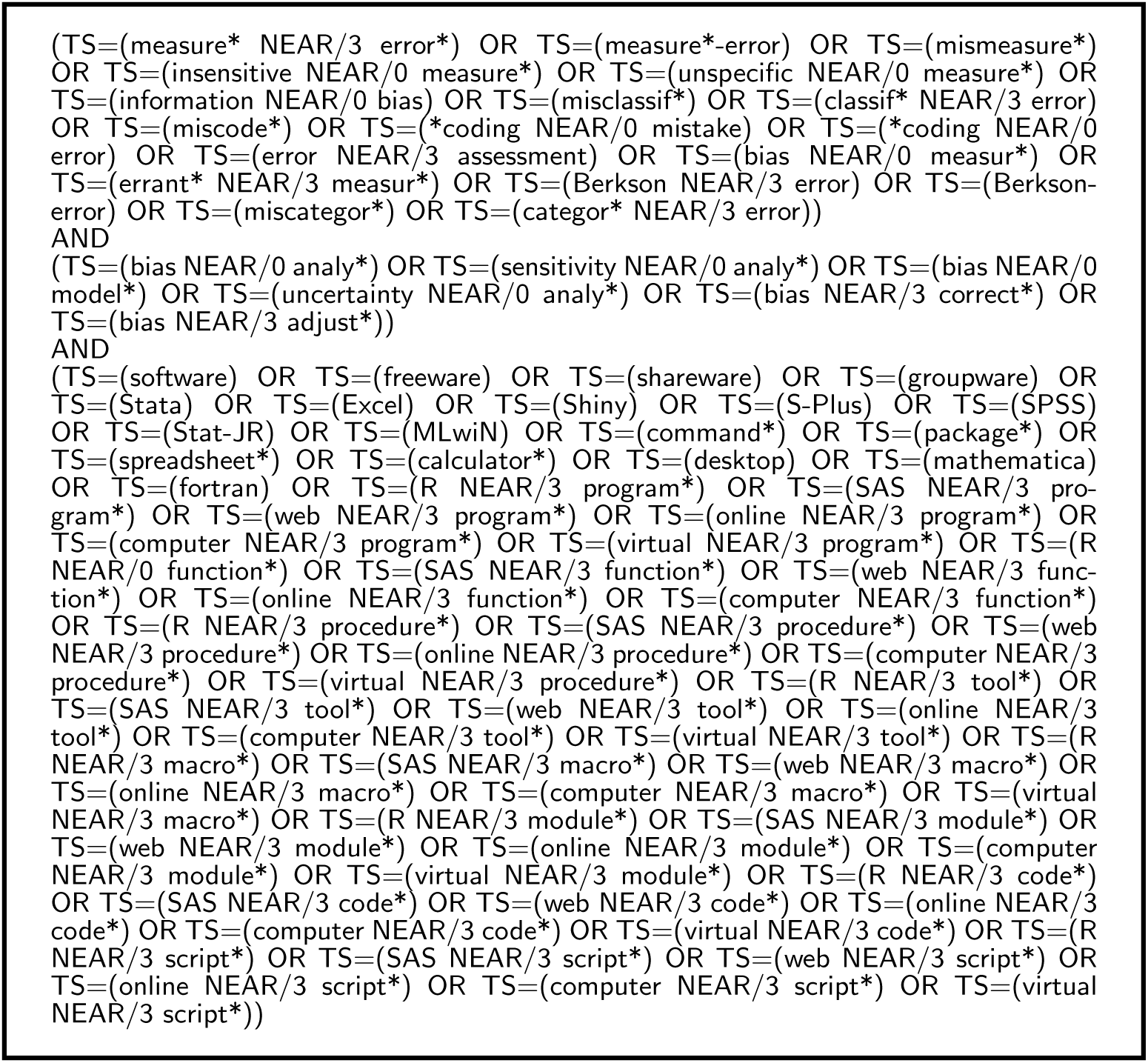
Clarviate Web of Science search terms for this software review [22].

**Fig. 2.**
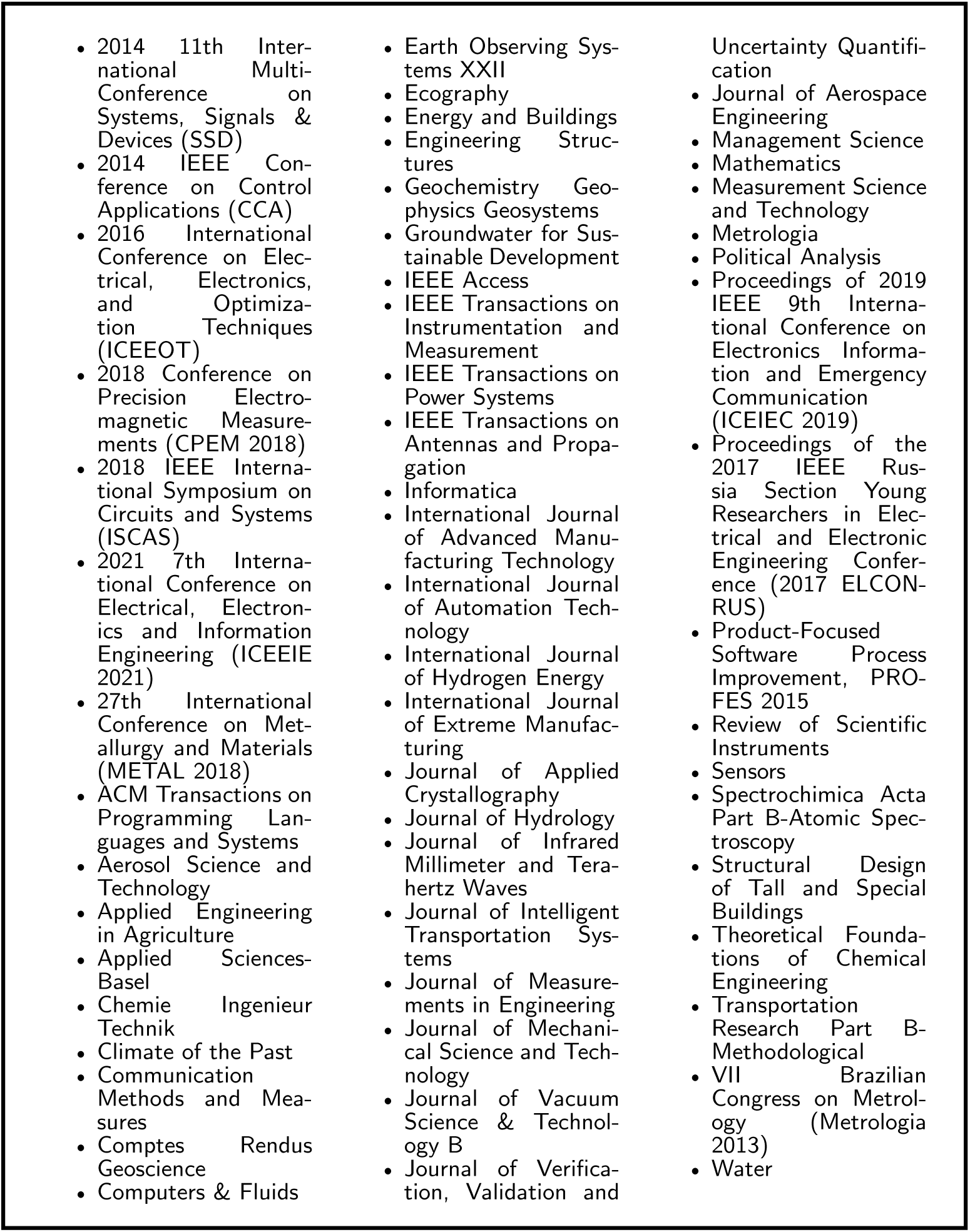
Journals outside of the scope of health research that were removed from our search results.

### 2.2 Software repository search

In order to capture software implementations not mentioned in published literature, we completed several additional searches outside of Web of Science. We searched The Comprehensive R Archive Network (CRAN) [23], R’s central software repository containing a large collection of quality-assured contributed packages. We also searched IDEAS/RePEc [24], an online database indexing items of economics research including articles as well as Boston College’s Statistical Software Components (SSC) archive, which contains user-written Stata commands and other code. In addition, using Stata’s *search* command, we conducted a search of the Stata manuals, the Stata Journal, and all Stata-related user-written commands that are available via Stata’s *net* command.

For our search of CRAN, we identified packages that mentioned both “measurement error” and “bias analysis” (or synonyms of these terms) in either their title or description. The R code used to implement this search is included in Additional File 2.

We first used R’s built-in CRAN package repository tools to extract the names, titles and descriptions of all of the packages maintained on CRAN on the search date, 8^th^ May 2024. After cleaning the extracted text, removing new line breaks and any multiple spaces, we then used the R *grep* function, which searches for pattern matches to its argument, to search for those packages which mentioned both “measurement error” and “bias analysis” in their title or description. Search terms and synonyms used were equivalent to those in Figure 1, in order to maintain consistency between our publication search and our repository search.

The “advanced search” tool of IDEAS was less flexible than the R functions used to search CRAN, and so for this database we simplified our search strategy. We searched IDEAS for software components that had both “measurement error” and “bias analysis” or their synonyms in any of their title, abstract or key words using the search string given in Figure 3. We also used this same set of terms to search the Stata manuals, the implementation of which can be found in Additional File 3. We considered all results which referenced either “measurement error” or “bias analysis” or their synonyms.

**Fig. 3.**
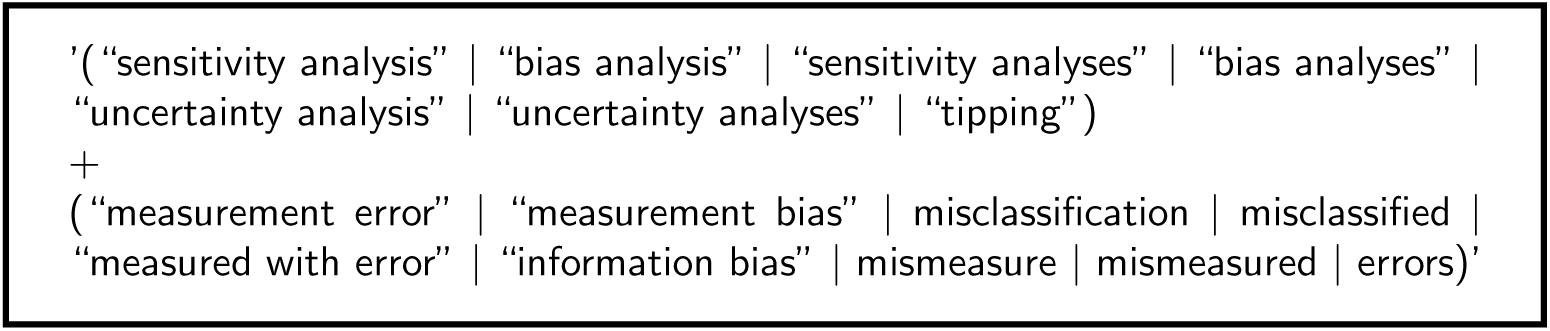
IDEAS/RePEc and Stata manual search terms for this software review.

We limited our results to tools which were first made publicly available (or had updated versions with new features implementing a QBA to mismeasurement) between 1^st^ January 2014 and the search date, 8^st^ May 2024 (inclusive).

### 2.3 Eligibility criteria

In our second step, the eligibility of identified abstracts and tools was assessed independently by two reviewers (CW and RH), with any disagreements resolved by consensus. Abstracts and tools were eligible for data extraction if they satisfied all of the following criteria:

1. the abstract mentioned purpose-built software,
2. the abstract discussed bias due to mismeasurement,
3. the software implemented a QBA for mismeasurement.

Examples of abstracts that would be excluded were those that only mentioned programming languages or code for examples rather than providing a purpose-built tool, abstracts where a QBA was not conducted but mentioned as further work, and abstracts which had software for purposes other than a QBA for mismeasurement.

### 2.4 Data extraction

In our third step, we examined the full texts of the included published papers and the documentation of the packages found via our CRAN, Stata manual, Stata *net* command and IDEAS/RePEc database searches to extract information about any software presented and its features. We excluded any R packages that had been removed from CRAN, software that could not be loaded, and software with example code that produced error messages. We also excluded any sensitivity analysis implementations that did not meet our definition for software or could not be considered a QBA due to not including at least one bias parameter.

Information collected included the year of development (or online publication) and the most recent recorded update. We also recorded the environment in which the tool was implemented (e.g., R, Stata, SAS, Excel, or web-based) and the types of outputs provided (tabular or graphical). To assess methodological capabilities, we examined the mismeasurement mechanisms accounted for (differential or non-differential) and whether multiple variables could be mismeasured simultaneously. We also categorised the type of data required (individual-level, aggregate counts, or summary statistics such as estimated regression coefficients) and the type of analysis supported. Further, we identified the variable affected by mismeasurement (outcome, exposure, or other covariates) and the nature of the observed variables (binary, categorical, or continuous). We also classified the type of QBA performed (deterministic or probabilistic) and, for deterministic QBAs, determined whether the tool supported a multidimensional bias analysis.

We also assessed the usability of each tool, firstly by whether usage examples were provided. Additionally, two reviewers (CW and RH) independently evaluated the level of detail in the documentation and the level of QBA knowledge required to perform a multidimensional or probabilistic QBA using the tool. Any disagreements were resolved by consensus.

Documentation was categorised into three levels; minimal, moderate and extensive. Tools classified as having minimal documentation provided only a brief description of the tool’s purpose, required inputs, and syntax (where applicable). Moderate documentation included a full description of each function, at least one usage example, a written explanation of the output, and a detailed description of the method implemented. Tools at this level also provided practice datasets where applicable. Tools classified as having extensive documentation offered additional tutorial materials such as vignettes, video tutorials, or an accompanying software journal article.

The level of QBA knowledge required was classified as either essential or specialist. Requiring essential knowledge indicated that the tool fully implemented a multidimensional or probabilistic bias analysis and displayed the results without users having to manually code these steps. Specialist knowledge was deemed to be required when users had to manually implement or visualize a multidimensional or probabilistic bias analysis. Alternatively, the tool may have required expertise in Bayesian methods, such as defining priors or assessing the convergence of MCMC samplers.

## 3 Results

After removal of duplicates, our initial Web of Science search returned 254 results. We then excluded 110 papers when restricting to publications made between 1^st^ January 2014 and 1^st^ May 2024. A further 63 papers were manually excluded that were published in journals outside of the scope of health research (as listed in Figure 2). We were left with a total of 81 abstracts. This initial search step is illustrated in Figure 4.

**Fig. 4.**
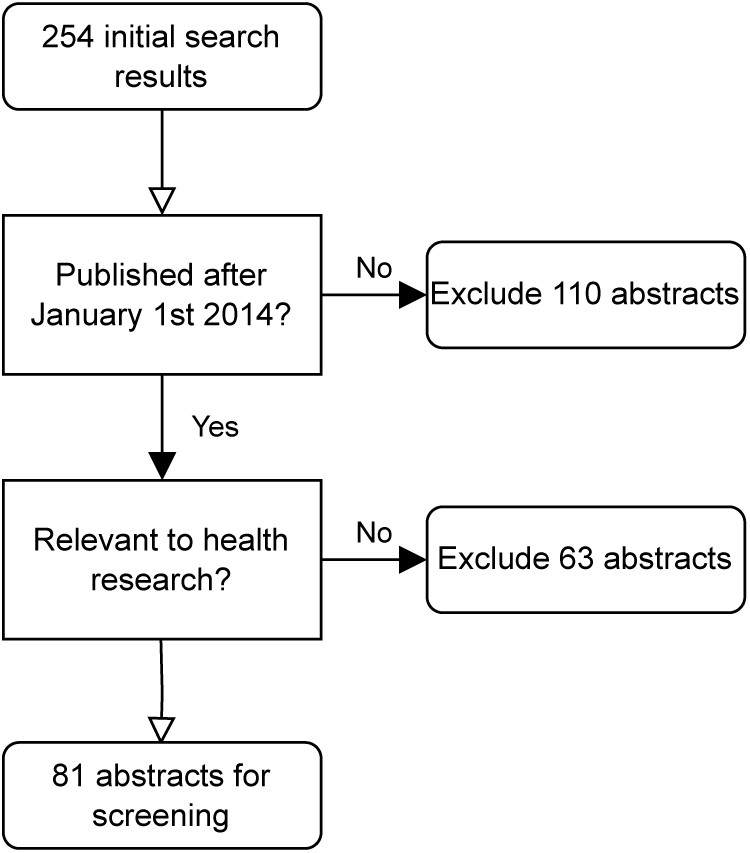
Flowchart of the publication search step of the review.

We excluded 37 abstracts which did not provide a purpose-built statistical software implementation, 10 abstracts that did not focus on bias due to mismeasurement, and nine abstracts where the software provided was not conducting a QBA for mismeasurement (e.g. the QBA was instead for an alternative form of bias). The abstract screening process is illustrated in Figure 5. When reviewing the full text of the remaining 25 articles, we found references to 24 unique software tools.

**Fig. 5.**
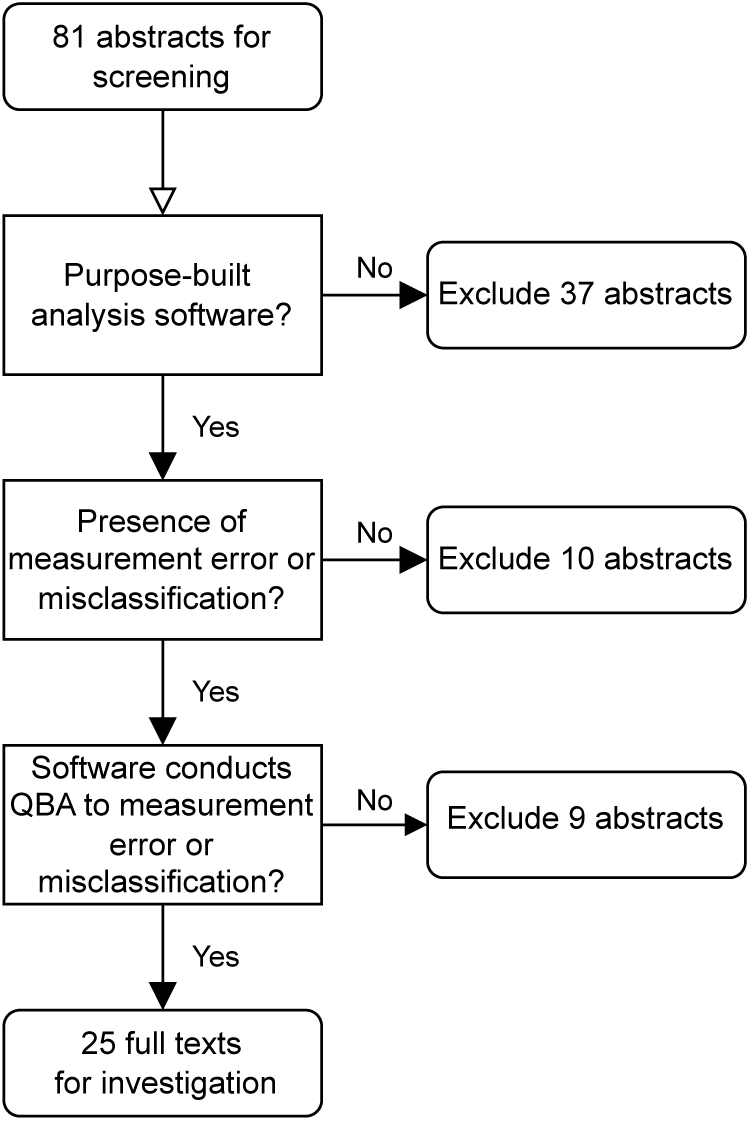
Flowchart of the abstract screening step of the review.

Our IDEAS search identified a single Stata command, *episens* [25, 26], however, the mismeasurement-related functions of this tool had not been updated since 2008, and so the tool fell outside of our date range for eligibility. We discuss this tool further in Section 3.1, in Table 2.

Our CRAN search returned ten software tools, four of which had also been identified by our Web of Science search. The tool *multibias* [27] also referenced an additional web tool implementation in its documentation, *multibias web tool* [28]. Thus, in total, our CRAN search provided an additional seven tools.

Our search using the Stata *search* command returned an initial 205 results, 136 of which were either duplicates or were outside of our date range and so were excluded. From the 69 remaining search results, four eligible tools were found.

In total, 35 unique software tools were identified across all of our searches, the process of which is summarised in Figure 6. Among these 35 tools, nine were excluded because they did not implement a QBA to mismeasurement, four were excluded because they did not provide sufficient documentation or were code for a specific example requiring user adjustment^1^, one tool was excluded because it was unable to run examples without errors at the time of review, and one tool was excluded because it had been removed from CRAN. The remaining 20 met our inclusion criteria.

**Fig. 6.**
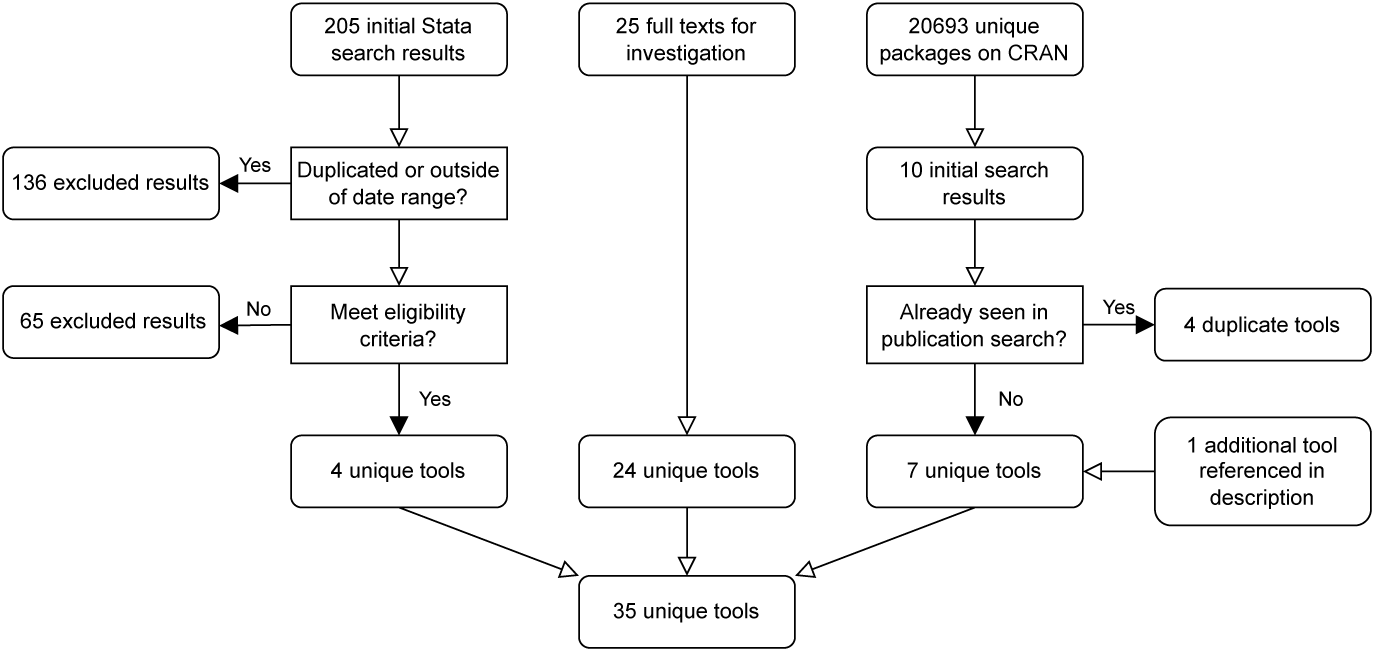
Flowchart of the software tool search process.

Of these 20 tools, *APScalculator* [31], *bamm* [32], and *SensiPhy* [33] do not implement a QBA for an exposure effect estimate of an epidemiological study and so are excluded from the results presented in Table 1. The web tool *APScalculator* evaluates the impact of classical measurement error [5, Chapter 1] on the categorization of a continuous variable into user-specified groups, rather than directly assessing bias in effect estimates. The R package *SensiPhy* estimates the impact of various sources of uncertainty in phylogenetic comparative methods used within ecology, which differs substantially from applications in health research. The Stata command *bamm* conducts a Bayesian bias analysis to investigate the distribution of a single misclassified categorical variable, which could be either nominal or ordinal.

**Table 1.** Software programs implementing a quantitative bias analysis for mismeasurement published between 2014 and 2024.

| Software name (Year) | Environment | Output |  | Mismeasurement |  |  | Bias analysis |  |
| --- | --- | --- | --- | --- | --- | --- | --- | --- |
|  |  | Table | Plot | Type | Differential | Multiple mismeasured variables | Type | Multi dimensional |
| *biasepi* (2019) [39, 40] | Stata command | ✓ | - | MC | D, ND | - | Det | - |
| *pvw* (2019) [41] | Stata command | ✓ | - | MC | D, ND | - | Det | - |
| *SAMBA-EHR* (2020) [42, 43] | Web tool | - | ✓ | MC | ND | - | Det | ✓ |
| *Outcome Misclassification* (2020) [44] | Web tool | ✓ | ✓ | MC | D, ND | - | Det | ✓ |
| *SensitivityAnalysis* (2020) [45] | Web tool | - | ✓ | MC | ND | - | Prob | - |
| *miCoPTCM* (2016, upd. 2020) [46] | R package | ✓ | - | ME | ND | ✓ | Det | - |
| *ivbounds* (2021) [47, 48] | Stata command | ✓ | - | MC | ND | - | Det | ✓ |
| *MediationSensitivityAnalysis* (2021) [49] | Web tool | ✓ | ✓ | ME | ND | ✓ | Det | ✓ |
| *BayesSenMC* (2021) [50] | R package | ✓ | ✓ | MC | D, ND | - | Prob | - |
| *EValue* (2017, upd. 2021) [51] | R package | ✓ | - | MC | D | ✓ | Det | - |
| *ConMed* (2023) [52] | R package:<br>non-CRAN | - | ✓ | ME | ND | ✓ | Det | - |
| *episensr* (2015, upd. 2023) [53] | R package | ✓ | ✓ | MC | D, ND | ✓ | Det, Prob | ✓ |
| *apisensr* (2021, upd. 2023) [54, 55] | Web tool | ✓ | ✓ | MC | D, ND | - | Det, Prob | ✓ |
| *mgee2* (2020, upd. 2023) [56, 57] | R package | ✓ | ✓ | MC | ND | ✓ | Det | - |
| *multibias web tool* (2023) [20, 28] | Web tool | ✓ | ✓ | MC | D, ND | - | Det, Prob | - |
| *multibias* (2023, upd. 2024) [20, 27] | R package | ✓ | - | MC | D, ND | ✓ | Det | - |
| *rcme* (2023, upd. 2024) [58] | R package:<br>non-CRAN | ✓ | ✓ | ME | D, ND | - | Det | - |

| Software name (Year) | Analysis |  |  |  |  |
| --- | --- | --- | --- | --- | --- |
|  | Data type | Analysis of interest | Outcome | Exposure | Other covariates |
| *biasepi* (2019) [39, 40] | Individual, Aggregate | Contingency table | **bin** | **bin** | - |
| *pvw* (2019) [41] | Individual | Regression | bin | **bin** | bin, cat, cts |
| *SAMBA-EHR* (2020) [42, 43] | Summary | Regression | **bin** | cts, cat | - |
| *Outcome Misclassification* (2020) [44] | Summary | Contingency table | **bin** | bin | - |
| *SensitivityAnalysis* (2020) [45] | Summary | Regression | cts | bin | **bin** |
| *miCoPTCM* (2016, upd. 2020) [46] | Individual | Survival analysis | TTE | **cts** | **cts** |
| *ivbounds* (2021) [47, 48] | Individual | Instrumental variable analysis | bin, cat, cts | **bin** | bin, cat, cts |
| *MediationSensitivityAnalysis* (2021) [49] | Summary | Mediation analysis | **cts** | bin | **Mediator (cts), other (cts)** |
| *BayesSenMC* (2021) [50] | Aggregate | Contingency table | bin | **bin** | - |
| *EValue* (2017, upd. 2021) [51, 59] | Summary | Regression | **bin** | **bin** | - |
| *ConMed* (2023) [52] | Summary | Mediation analysis | **cts** | cts | **Mediator (cts), other (bin, cat, cts)** |
| *episensr* (2015, upd. 2023) [53] | Aggregate | Contingency table | **bin, TTE** | **bin** | **bin** |
| *apisensr* (2021, upd. 2023) [55] | Aggregate | Contingency table | **bin** | **bin** | **bin** |
| *mgee2* (2020, upd. 2023) [56, 57] | Individual | Longitudinal analysis | **cat (ordinal)** | **cat (ordinal),**<br>bin, cts | **cat (ordinal),**<br>bin, cat, cts |
| *multibias web tool* (2023) [20, 28] | Individual | Regression | bin | **bin** | bin, cat, cts |
| *multibias* (2023, upd. 2024) [20, 27] | Individual | Regression | **bin** | **bin** | bin, cat, cts |
| *rcme* (2023, upd. 2024) [58] | Individual | Regression | **cts** | **cts** | **cts** |

| Software name (Year) | Level of documentation | Usage |  |
| --- | --- | --- | --- |
|  |  | Examples | QBA knowledge required |
| *biasepi* (2019) [39, 40] | Moderate | ✓ | Specialist |
| *pvw* (2019) [41] | Moderate | ✓ | Specialist |
| *SAMBA-EHR* (2020) [42, 43] | Extensive | ✓ | Essential |
| *Outcome Misclassification* (2020) [44] | Extensive | ✓ | Essential |
| *SensitivityAnalysis* (2020) [45] | Moderate | ✓ | Essential |
| *miCoPTCM* (2016, upd. 2020) [46] | Moderate | ✓ | Specialist |
| *ivbounds* (2021) [47, 48] | Extensive | ✓ | Essential |
| *MediationSensitivityAnalysis* (2021) [49] | Minimal | - | Essential |
| *BayesSenMC* (2021) [50] | Extensive | ✓ | Specialist |
| *EValue* (2017, upd. 2021) [51] | Extensive | ✓ | Essential |
| *ConMed* (2023) [52] | Moderate | ✓ | Essential |
| *episensr* (2015, upd. 2023) [53] | Extensive | ✓ | Essential |
| *apisensr* (2021, upd. 2023) [55] | Extensive | ✓ | Essential |
| *mgee2* (2020, upd. 2023) [56, 57] | Moderate | ✓ | Specialist |
| *multibias web tool* (2023) [20, 28] | Minimal | - | Essential |
| *multibias* (2023, upd. 2024) [20, 27] | Extensive | ✓ | Specialist |
| *rcme* (2023, upd. 2024) [58] | Minimal | ✓ | Specialist |

Abbreviations used (in alphabetical order): upd.; Updated.

Table 1 summarises the key features of the 17 total software programs we found that are applicable to health studies aiming to quantify bias in an effect estimate, in order of most recent update. Of the tools reviewed, eight (47%) are implemented as R packages, six (35%) are web-based applications, and three (18%) are Stata commands.^2^

Of the 17 tools, four implement a QBA in the case of measurement error in a continuous variable (*miCoPTCM*, *MediationSensitivityAnalysis*, *ConMed* and *rcme*). Only *rcme* allows for multiplicative measurement error (i.e. error that scales with the true value of the variable), whilst the rest employ a classical additive measurement error model [5, Chapter 1]. A total of 12 tools apply a QBA for misclassification of a binary variable. Only one tool, *mgee2*, implements a QBA when the misclassified variable has more than two categories.

In total, 11 software tools handle cases of outcome mismeasurement, 12 handle exposure mismeasurement, and seven handle mismeasurement in other covariates (such as effect modifiers, mediators, or potential confounders). Among these, two tools (*SAMBA-EHR* and *Outcome Misclassification*) focus solely on outcome mismeasurement, four (*pvw*, *ivbounds*, *BayesSenMC* and *multibias web tool*) exclusively handle exposure mismeasurement, and only *SensitivityAnalysis* is specific to misclassification of a confounder.

All tools except *biasepi*, which can take either individual or aggregate data as inputs, are specific in the data type required. Individual-level data is required by seven of the tools, aggregated count data by four tools, and summary statistics (such as regression coefficients or other statistics derived from the data) are required by six of the tools.

When the analysis of interest is a mediation analysis, two tools are applicable: *MediationSensitivityAnalysis* and *ConMed*. *MediationSensitivityAnalysis* performs bias analysis for measurement error in the outcome, mediator, or other observed covariates (effect modifiers or potential confounders), with the assumption that the binary exposure is measured without error. *ConMed* adjusts for measurement error in the mediator or outcome, and is applicable specifically when there is unmeasured confounding as well as measurement error.

For other types of analysis of interest, R package *miCoPTCM* accounts for measurement error of a continuous covariate or exposure in survival analysis (specifically a promotion time cure model), where the outcome is a time-to-event variable. Stata command *ivbounds* applies QBA for an instrumental variable analysis, allowing for a binary or categorical instrumental variable. R package *mgee2* conducts QBA for a longitudinal analysis, where there are changes within the same individuals or groups over time. Of the remaining tools, five are applicable for the analysis of contingency tables and seven for logistic or linear regression.

Most tools require the outcome variable of the analysis of interest to be either binary (nine programs) or continuous (five programs). Only *mgee2* and *ivbounds* allow for a discrete outcome variable with more than two categories. The exposure variable of the analysis of interest is typically required to be exclusively either binary (nine programs) or continuous (four programs), but two programs (*SAMBA-EHR* and *mgee2*) allow exposure variables to be of multiple types including both discrete or continuous options. Of all of the tools, 12 (71%) allow for the inclusion of other covariates than just the exposure and outcome in the analysis.

Among the tools, nine are applicable for both differential and non-differential mismeasurement, while seven are for non-differential mismeasurement only. The tool *EValue* is specific to differential misclassification. Multiple variables can be mismeasured simultaneously in seven (41%) of the tools.

A deterministic QBA is implemented exclusively in 11 tools, two (*SensitivityAnalysis* and *BayesSensMC*) support only a probabilistic QBA, and three include options to implement both a deterministic and probabilistic QBA (*episensr*, *apisensr*, and *multibias web tool* ^3^). Among the tools that implement a probabilistic QBA, only *BayesSenMC* performs a Bayesian bias analysis, and the remaining tools perform a Monte Carlo bias analysis. Among the tools that implement a deterministic QBA, only six (40%) perform a multidimensional analysis.

Approximately half of the tools (eight) provide both graphical and tabular outputs to aid the user in interpreting the results. Only three tools do not produce tables: *SAMBA-EHR*, *SensitivityAnalysis* and *ConMed*. Three R packages and three Stata commands do not provide graphical plots of their results.

Tools *biasepi*, *multibias*, *multibias web tool*, *ConMed*, *EValue*, *episensr* and *Medi-ationSensitivityAnalysis* can also perform a multiple QBA for mismeasurement and unmeasured confounding, with *biasepi*, *EValue*, *multibias*, and *multibias web tool* also able to adjust for selection bias. Note that multiple sources of bias are simultaneously adjusted by all tools except *EValue*, *biasepi* and *episensr*, which instead use a sequential approach.

Of the 17 tools reviewed, only two do not include usage examples, both of which are web tools. Documentation quality varies: eight tools have extensive documentation, six have moderate documentation, and three have minimal documentation. Ten tools fully implement a multidimensional or probabilistic QBA (i.e., user only requires essential QBA knowledge as the software implements all steps of the QBA including summaries of the results). However, seven tools require users to have specialist knowledge (e.g., a tool only performs a simple bias analysis and so a user must write their own code to conduct a probabilistic QBA using this tool).

### 3.1 Grey literature

In our formal search we focused on software described in the published literature between January 1^st^ 2014 and May 1^st^ 2024, or software made available during this period via CRAN, the Stata manuals, the IDEAS/RePEc database, or other Stata user-written commands available using Stata’s *net* command. This approach ensured we captured the latest tools which applied researchers could readily locate. However, we recognise that additional software exists that was not identified through this search. For example, some tools have not been mentioned in journal articles and are hosted in alternative environments such as web-based platforms. Others were developed before 2014 and have not been significantly updated since but remain in use. Table 2 gives a non-exhaustive brief overview of some tools known to the authors that were not captured by our formal search strategy, but that may be of interest to researchers. *Short code* [29, 30], which was excluded from our formal search results due to not having adequate documentation (see Section 3), is also included in Table 2 because of its prominence in the QBA literature and potential use alongside [11] for those learning to conduct a QBA.

**Table 2.** Software tools for mismeasurement correction and QBA not captured by our formal search strategy.

| Tool | Environment | Brief description |
| --- | --- | --- |
| *Quantitative Bias Analysis* [60] | Web tool | Conducts simple and multidimensional bias analyses for misclassification and unmeasured confounding |
| *CMAverse* [61, 62] | R package (non-CRAN) | Conducts sensitivity analyses for unmeasured confounding, measurement error, and selection bias in causal mediation analysis. |
| *episens* [25, 26] | Stata | Provides basic sensitivity analysis of the observed relative risks, adjusting for unmeasured confounding and misclassification of the exposure |
| *eivtools* [63] | R package | Functions for analysis with error-prone covariates |
| *Prob Bias Analysis for Information Bias* [64] | Web tool | Probabilistic bias analysis for misclassification; an R based web interface for *episensr* [53]. |
| *simplemba* [65] | Web tool | Odds ratio calculator with misclassification, selection bias, and confounding adjustment. |
| *sensmac* [66, 67] | SAS | Implements probabilistic sensitivity analysis to misclassification of a binary variable. |
| *Multiple bias model* [68, 69] | SAS | Code and dataset for conducting multiple bias modelling. |
| *Mecor* [70] | R package | Functions to perform covariate measurement error correction. |
| *Short code* [29, 30] | R and SAS code | Code to perform record and summary level QBA, for misclassification and confounding. |

## 4 Discussion

We have conducted an up-to-date review of software implementations of QBA to mis-measurement described in the published literature, R packages available on CRAN, and Stata commands, including user-written commands available from the SSC archive or via Stata’s *net* command. All software tools were developed or significantly updated post-2019, with most (65%) having been developed or updated since 2021. The software tools were either R packages, Stata commands or online web tools and were available for routine analyses of interest such as linear regression and contingency tables, and for more specialized analyses such as mediation analysis, instrumental variable analysis and survival analysis. All but one software tool implemented a QBA to non-differential mismeasurement with just over half applicable for differential and non-differential mismeasurement. Also, more than half of the software tools implement a QBA for misclassification of a binary variable. Although most software tools implemented a deterministic bias analysis, only six (40%) of these tools included features to allow the user to perform a multidimensional bias analysis. Most tools provided usage examples, but documentation quality varied from minimal to extensive. While several tools offered comprehensive guidance, including tutorials and vignettes, others provided only brief descriptions of inputs and outputs. Just over half of the tools implemented all the steps of a multidimensional or probabilistic QBA for the user, but a subset required specialist QBA knowledge, such as understanding Bayesian priors or manually implementing a multidimensional bias analysis.

Although previous work has suggested that implementation challenges in QBA have largely been addressed [18], our findings indicate significant gaps remain. Existing tools rarely support QBA for categorical variables with more than two levels, and few address measurement error in continuous variables beyond classical error models. Further, many of the tools reviewed were designed for specific use cases, often allowing only a single data type or a single type of bias analysis. Developing or expanding software to cover these scenarios, and to handle multiple potential mismeasurement types, would improve accessibility and could lead to greater uptake by applied researchers.

In addition to gaps in available methods, documentation inconsistencies pose a barrier to effective tool use. Although all tools met our definition of “software” by including documentation, we found that documentation often did not explicitly state key assumptions about the data. For example, despite eight tools allowing multiple variables to be simultaneously measured with error, only *episensr*explicitly stated that errors were required to be independent. The lack of clear statements about underlying assumptions forces users to rely on prior methodological knowledge or manual code inspection, increasing the risk of misuse or misinterpretation. This not only exacerbates the broader issue of unacknowledged dependent error in epidemiological studies [38] but also creates a barrier to the adoption of tools by applied researchers. Addressing these gaps by explicitly stating assumptions and tool limitations in documentation could facilitate wider adoption and correct application of QBA methods. Future work could enhance this review by providing applied examples of the usage of tools with simulated or real data, clarifying when each tool is most appropriate.

A limitation of our work is that we restricted our search to epidemiology, statistics, and health journals. This may have excluded software from other disciplines that could be applicable to health research. The numbers of these tools would likely be small, however, as different fields face very different complications and considerations to medical research. Further, it is unlikely that health researchers would look outside of their field in order to find tools for use. Conducting similar reviews in domains such as psychology, engineering, or computational biology could increase awareness of potentially useful tools across disciplines.

The substantial number of tools in Table 2 that were not identified through our formal search highlights the challenge for applied researchers in discovering relevant QBA software. Many of these tools would be difficult for researchers unfamiliar with mismeasurement and quantitative bias analysis to discover, reinforcing the need for greater visibility of tools. Publishing software tools in widely recognized repositories, maintaining clear documentation of updates and expansions, and encouraging researchers to cite software in their outputs would help bridge this gap.

## 5 Conclusions

Our review highlights some progress in the availability of software tools for QBA to mismeasurement but also reveals important gaps that limit their accessibility and applicability. While many tools support common analyses and provide extensive documentation, others lack clarity on key assumptions, require specialist knowledge, or are restricted to specific use cases. There is a lack of tools for handling misclassification of categorical variables and for addressing non-classical measurement error in continuous variables. Improved documentation, broader methodological coverage, and increased visibility through publication in software journals and good citation practices could enhance the usability and adoption of these tools. Future efforts should focus on developing more comprehensive software tools and ensuring that researchers can easily identify and apply appropriate programs for addressing mismeasurement in their studies.

## Supporting information

Additional files as detailed in manuscript

## Data Availability

This study did not involve any underlying
data. Computing code is available as supplemental digital content.

## Declarations

### Ethics approval and consent to participate

Not applicable.

### Consent for publication

Not applicable.

### Availability of data and materials

This study did not involve any underlying data. Computing code available as supplemental digital content. All software programs are freely available as detailed in their documentation (see references).

### Abbreviations

bin: Binary
cat: Categorical
cts: Continuous
D: Differential
Det: Deterministic
MC: Misclassification
ME: Measurement error
ND: Non-differential
Prob: Probabilistic
QBA: Quantitative bias analysis;
TTE: Time-to-event
upd.: Updated.

### Authors’ contributions

RAH proposed and designed the study. CJCW carried out the statistical analyses with participation from RAH. CJCW, RAH, KT, and JWB drafted the manuscript. All authors read and approved the final version of the manuscript.

## Additional files

### Additional file 1

**Format:** .pdf

**Title:** PRISMA-ScR Checklist

**Description:** Filled PRISMA-ScR checklist for our scoping review methodology.

### Additional file 2

Format: .R

**Title:** CRAN Search Code

**Description:** R code used to conduct the CRAN search stage.

### Additional file 3

**Format:** .do

**Title:** Stata Search Code

**Description:** Stata code used to conduct the search of the Stata manual, Stata journal and user written code.

### Table 1 file

**Format:** .xls

**Title:** Software programs implementing a quantitative bias analysis for mismeasurement published between 2014 and 2024.

**Description:** An excel spreadsheet containing Table 1, also included in .tex form in this manuscript file. Table caption and footnotes are included in the file.

## Footnotes

1 One of these tools, *Short code* [29, 30], is discussed further in Section 3.1 and included in Table 2, due to its prominence in core QBA literature [11].

2 We note here that the R package *EValue* has a corresponding Stata implementation *evalue* [34] and web tool [35, 36]. At the time of writing, neither of these tools implement a QBA to mismeasured variables; instead, they focus on bias due to unmeasured confounding, so we have not included them here. See [12] for further details on these tools for QBA to unmeasured confounding. The web tool *SAMBA-EHR* also exists alongside R package *SAMBA* [37]. This R package does not implement a QBA to mismeasured variables and so has not been reviewed here.

3 The tool multibias does not perform a probabilistic QBA, but does provide guidance and example code demonstrating how a user can manually implement a probabilistic QBA using the tool. The multibias web tool, however, provides a function which performs a probabilistic QBA.

